# Automatic Head and Neck Tumor Segmentation in PET/CT with Scale Attention Network

**DOI:** 10.1101/2020.11.11.20230185

**Authors:** Yading Yuan

**Affiliations:** Department of Radiation Oncology, Icahn School of Medicine at Mount Sinai, New York, NY, USA

## Abstract

Automatic segmentation is an essential but challenging step for extracting quantitative imaging bio-markers for characterizing head and neck tumor in tumor detection, diagnosis, prognosis, treatment planning and assessment. The HEad and neCK TumOR Segmentation Challenge 2020 (HECKTOR 2020) provides a common platform for comparing different automatic algorithms for segmentation the primary gross target volume (GTV) in the oropharynx region on FDG-PET and CT images. We participated in the image segmentation challenge by developing a fully automatic segmentation network based on encoder-decoder architecture. In order to better integrate information across different scales, we proposed a dynamic scale attention mechanism that incorporates low-level details with high-level semantics from feature maps at different scales. Our framework was trained using the 201 challenge training cases provided by HECKTOR 2020, and achieved an average Dice Similarity Coefficient (DSC) of 0.7505 with cross validation. By testing on the 53 testing cases, our model achieved an average DSC, precision and recall of 0.7318, 0.7851, and 0.7319 respectively, which ranked our method in the fourth place in the challenge (id: deepX).

## 1 Introduction

Head and Neck (H&N) cancers are among the most common cancers worldwide (the 5th leading cancer by incidence) [1]. Radiotherapy (RT) combined with chemotherapy is the standard treatment for patients with inoperable H&N cancers [2]. However, studies showed that locoregional failures occur in up to 40% of patients in the first two years after the treatment [3]. In order to identify patients with a worse prognosis before treatment, several radiomic studies have been recently proposed to leverage the massive quantitative features extracted from high-dimensional imaging data acquired during diagnosis and treatment. While these studies showed promising results, their generalization performance needs to be further validated on large patient cohorts. However, the primary tumors and nodal metastases are currently delineated by oncologists by reviewing both PET and CT images simultaneously, which is impractical and error-prone when scaling up to a massive patient population. In addition, radiation oncologists also need to manually delineate the treatment targets and the organs at risk (OARs) when designing a treatment plan for radiotherapy, which is time-consuming and suffers from inter- and intra-operator variations [4]. As a result, automated segmentation methods have been of great demand to assist clinicians for better detection, diagnosis, prognosis, treatment planing as well as assessment of H&N cancers.

The HEad and neCK TumOR segmentation challenge (HECKTOR) [5, 6] aims to accelerate the research and development of reliable methods for automatic H&N primary tumor segmentation on oropharyngeal cancers by providing a large PET/CT dataset that includes 201 cases for model training and 53 cases for testing, as an example shown in Fig. 1. For training cases, the ground truth was annotated by multiple radiation oncologists, either directly on the CT of the PET/CT study (31% of the patients) or on a different CT scan dedicated to treatment planning (69%) where the planning CT was registered to the PET/CT scans [7]. While the testing cases were directly annotated on the PET/CT images. The cases were collected from five different institutions where four of them (CHGJ, CHMR, CHUM and CHUS) will be used for training and the remaining one (CHUV) will be used for testing. Each case includes a co-registered PET-CT set as well as the primary Gross Tumor Volume (GTVt) annotation. A bounding box was also provided to enable the segmentation algorithms focus on the volume of interest (VOI) near GTVt [8]. These images were resampled to 1×1×1 mm isotropic resolution and then cropped to a volume size of 144×144×144. The evaluation will be based on Dice Similarity Coefficient (DSC), which is computed only within these bounding boxes at the original CT resolution.

**Fig. 1.**
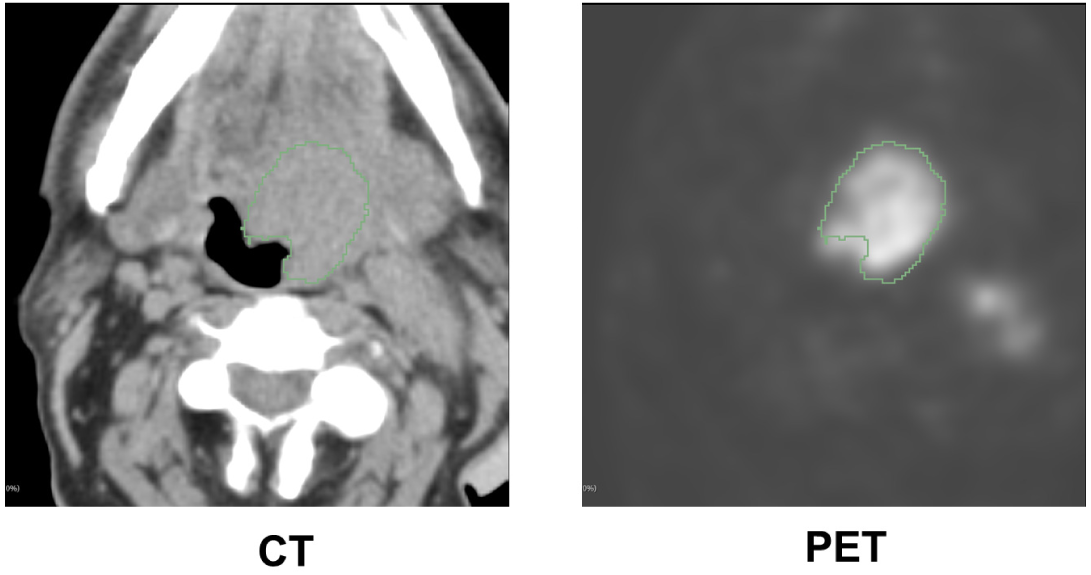
An example of PET/CT used in HECKTOR 2020 challenge

### 2 Related work

While convolutional neural networks have been successfully applied in various biomedical image segmentation tasks, only few studies have been conducted in the applications of deep convolutional neural networks (DCNNs) in automated segmentation of tumors in PET/CT images. In [9], Moe et al. presented a PET-CT segmentation algorithm based on 2D U-Net architecture to delineate the primary tumor as well as metastatic lymph nodes. Their model was trained with 152 patients and tested on 40 patients. Andrearczyk et al. [5] expanded this work by investigating several segmentation strategies based on V-Net architecture on a publicly available dataset with 202 patients. Zhao et al. [10] employed a multi-modality fully convolutional network (FCN) for tumor co-segmentation in PET-CT images on a clinic dataset of 84 patients with lung cancer, and Zhong et al. [11] proposed a segmentation method that consists of two coupled 3D U-Nets for simultaneously co-segmenting tumors in PET/CT images for 60 non-small cell lung cancer (NSCLC) patients.

The success of U-Net [13] and its variants in automatic PET-CT segmentation is largely contributed to the skip connection design that allows high resolution features in the encoding pathway be used as additional inputs to the convolutional layers in the decoding pathway, and thus recovers fine details for image segmentation. While intuitive, the current U-Net architecture restricts the feature fusion at the same scale when multiple scale feature maps are available in the encoding pathway. Studies have shown feature maps in different scales usually carry distinctive information in that low-level features represent detailed spatial information while high-level features capture semantic information such as target position, therefore, the full-scale information may not be fully employed with the scale-wise feature fusion in the current U-Net architecture.

To make full use of the multi-scale information, we propose a novel encoder-decoder network architecture named scale attention networks (SA-Net), where we re-design the inter-connections between the encoding and decoding pathways by replacing the scale-wise skip connections in U-Net with full-scale skip connections. This allows SA-Net to incorporate low-level fine details with the high-level semantic information into a unified framework. In order to highlight the important scales, we introduce the attention mechanism [14, 15] into SA-Net such that when the model learns, the weight on each scale for each feature channel will be adaptively tuned to emphasize the important scales while suppressing the less important ones. Figure 2 shows the overall architecture of SA-Net.

**Fig. 2.**
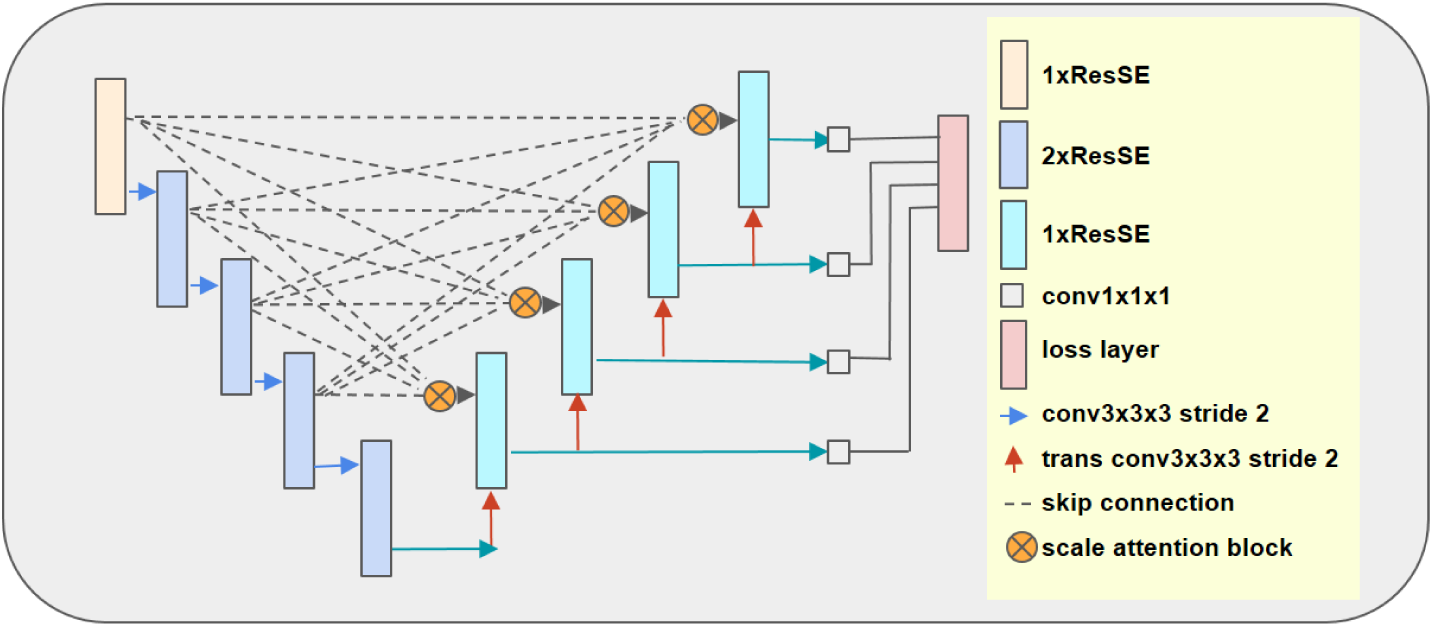
Architecture of SA-Net. Input is a 2 × 128 × 128 × 128 tensor followed by one ResSE block with 24 filters. Here ResSE stands for a squeeze-and-excitation block embedded in a residual module [14]. By progressively halving the feature map dimension while doubling the feature width at each scale, the endpoint of the encoding pathway has a dimension of 384 × 8 × 8 × 8. The output of the encoding pathway has one channel with the same spatial size as the input, i.e., 1 × 128 × 128 × 128.

## 3 Methods

### 3.1 Overall network structure

SA-Net follows a typical encoding-decoding architecture with an asymmetrically larger encoding pathway to learn representative features and a smaller decoding pathway to recover the segmentation mask in the original resolution. The outputs of encoding blocks at different scales are merged to the scale attention blocks (SA-block) to learn and select features with full-scale information. Due to the limit of GPU memory, we convert the input image from 144 × 144 × 144 to 128 × 128 × 128, and concatenate PET and CT images of each patient into a two channel tensor to yield an input to SA-Net with the dimension of 2 × 128 × 128×128. The network output is a map with size of 1 × 128 ×128 × 128 where each voxel value represents the probability that the corresponding voxel belongs to the tumor target.

### 3.2 Encoding pathway

The encoding pathway is built upon ResNet [16] blocks, where each block consists of two Convolution-Normalization-ReLU layers followed by additive identity skip connection. We keep the batch size to 1 in our study to allocate more GPU memory resource to the depth and width of the model, therefore, we use instance normalization, i.e., group normalization [21] with one feature channel in each group, which has been demonstrated with better performance than batch normalization when batch size is small. In order to further improve the representative capability of the model, we add a squeeze-and-excitation module [14] into each residual block with reduction ratio *r* = 4 to form a ResSE block. The initial scale includes one ResSE block with the initial number of features (width) of 24. We then progressively halve the feature map dimension while doubling the feature width using a strided (stride=2) convolution at the first convolution layer of the first ResSE block in the adjacent scale level. All the remaining scales include two ResSE blocks and the endpoint of the encoding pathway has a dimension of 384 × 8 × 8 × 8.

### 3.3 Decoding pathway

The decoding pathway follows the reverse pattern of the encoding one, but with a single ResSE block in each spatial scale. At the beginning of each scale, we use a transpose convolution with stride of 2 to double the feature map dimension and reduce the feature width by 2. The upsampled feature maps are then added to the output of SA-block. Here we use summation instead of concatenation for information fusion between the encoding and decoding pathways to reduce GPU memory consumption and facilitate the information flowing. The endpoint of the decoding pathway has the same spatial dimension as the original input tensor and its feature width is reduced to 1 after a 1×1×1 convolution and a sigmoid function.

In order to regularize the model training and enforce the low- and middle-level blocks to learn discriminative features, we introduce deep supervision at each intermediate scale level of the decoding pathway. Each deep supervision subnet employs a 1 × 1 × 1 convolution for feature width reduction, followed by a trilinear upsampling layer such that they have the same spatial dimension as the output, then applies a sigmoid function to obtain extra dense predictions. These deep supervision subnets are directly connected to the loss function in order to further improve gradient flow propagation.

### 3.4 Scale attention block

The proposed scale attention block consists of full-scale skip connections from the encoding pathway to the decoding pathway, where each decoding layer incorporates the output feature maps from all the encoding layers to capture fine-grained details and coarse-grained semantics simultaneously in full scales. As an example illustrated in Fig. 3, the first stage of the SA-block is to add the input feature maps at different scales from the encoding pathway, represented as {*S*_*e*_, *e* = 1, …, *N*} where *N* is the number of total scales in the encoding pathway except the last block (*N* = 4 in this work), after transforming them to the feature maps with the same dimensions, i.e., *S*_*d*_ = Σ *f*_*ed*_(*S*_*e*_). Here *e* and *d* are the scale level at the encoding and decoding pathways, respectively. The transform function *f*_*ed*_(*S*_*e*_) is determined as follows. If *e < d, f*_*ed*_(*S*_*e*_) downsamples *S*_*e*_ by 2^(*d*−*e*)^ times by maxpooling followed by a Conv-Norm-ReLU block; if *e* = *d, f*_*ed*_(*S*_*e*_) = *S*_*e*_; and if *e > d, f*_*ed*_(*S*_*e*_) upsamples *S*_*e*_ through tri-linear upsamping after a Conv-Norm-ReLU block for channel number adjustment. For *S*_*d*_, a spatial pooling is used to average each feature to form an information embedding tensor 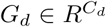, where *C*_*d*_ is the number of feature channels in scale *d*. Then a 1 −*to* −*N* Squeeze-Excitation is performed in which the global feature embedding *G*_*d*_ is squeezed to a compact feature 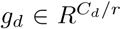 by passing through a fully connected layer with a reduction ratio of *r*, then another *N* fully connected layers with sigmoid function are applied for each scale excitation to recalibrate the feature channels on that scale. Finally, the contribution of each scale in each feature channel is normalized with a softmax function, yielding a scale-specific weight vector for each channel as 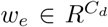, and the final output of the scale attention block is 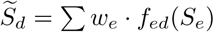.

**Fig. 3.**
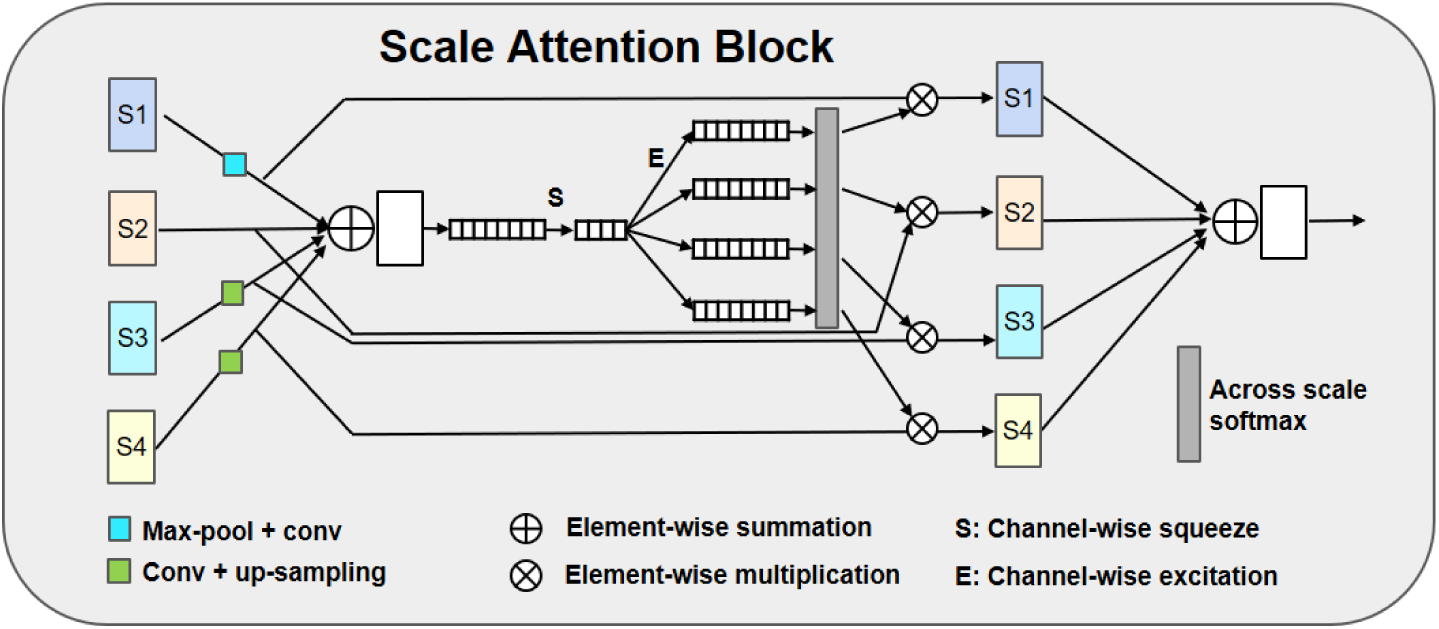
Scale attention block. Here *S*1, *S*2, *S*3 and *S*4 represent the input feature maps at different scales from the encoding pathway.

### 3.5 Implementation

Our framework was implemented with Python using Pytorch package. All the following steps were performed on the volumes of interest (VOIs) within the given bounding boxes. As for pre-processing, we truncated the CT numbers to [-125, 225] HU to eliminate the irrelevant information, then normalized the CT images with the mean and standard deviation of the HU values within GTVs in the entire training dataset. For PET images, we simply normalized each patient independently by subtracting the mean and dividing by the standard deviation of the image within the body. The model was trained with a patch size of 128×128×128 voxels and batch size of 1. We used Jaccard distance, which we developed in our previous studies [17–20], as the loss function in this work. It is defined as:

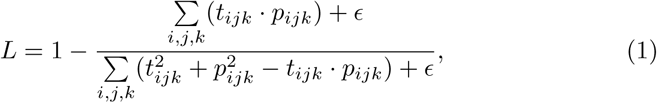

where *t*_*ijk*_ ∈ *{*0, 1*}* is the actual class of a voxel***x***_*ijk*_ with *t*_*ijk*_ = 1 for tumor and *t*_*ijk*_ = 0 for background, and *p*_*ijk*_ is the corresponding output from SA-Net. *∈* is used to ensure the stability of numerical computations.

Training the entire network took 300 iterations from scratch using Adam stochastic optimization method. The initial learning rate was set to 0.003, and learning rate decay and early stopping strategies were utilized when validation loss stopped decreasing. In particular, we kept monitoring the validation loss (*L*^(*valid*)^) in each iteration. We kept the learning rate unchanged at the first 150 iterations, but dropped the learning rate by a factor of 0.3 when *L*^(*valid*)^ stopped improving within the last 30 iterations. The model that yielded the best *L*^(*valid*)^was recorded for model inference.

In order to reduce overfitting, we randomly flipped the input volume in left/right, superior/inferior, and anterior/posterior directions on the fly with a probability of 0.5 for data augmentation. Other geometric augmentations included rotating input images by a random angle between [-10, 10] degrees and scaling them by a factor randomly selected from [0.9, 1.1]. We also adjusted the contrast in each image input channel by a factor randomly selected from [0.9, 1.1]. We used 5-fold cross validation to evaluate the performance of our model on the training dataset, in which a few hyper-parameters such as the feature width and input dimension were also experimentally determined. All the experiments were conducted on Nvidia GTX 1080 TI GPU with 11 GB memory.

We employed two different strategies to convert the 144 × 144 × 144 VOIs into 128 × 128 × 128 patches. In the first approach, we simply resized the original VOIs during the training and testing phases, in which image data were resampled isotropically using linear interpolation while the binary mark resampled with near neighbor interpolation. In the other approach, we randomly extracted a patch with a size of 128×128×128 from the VOIs during the training, and applied the sliding window to extract 8 patches from the VOI (2 windows in each dimension) and averaged the model outputs in the overlapping regions before applying a threshold of 0.5 to obtain a binary mask.

## 4 Results

We trained SA-Net with the training set provided by the HECKTOR 2020 challenge, and evaluated its performance on the training set via 5-fold cross validation. Table 1 shows the segmentation results in terms of DSC for each fold. As compared to the results in table 2 that were obtained from a model using the standard U-Net skip connections, the proposed SA-Net improved segmentation performance by 3.2% in patching and 1.3% in resizing, respectively.

**Table 1.** Segmentation results (DSC) of SA-Net in 5-fold cross validation using the training image dataset.

|  | fold-1 | fold-2 | fold-3 | fold-4 | fold-5 | Average |
| --- | --- | --- | --- | --- | --- | --- |
| Patching | 0.772 | 0.722 | 0.751 | 0.759 | 0.743 | 0.749 |
| Resizing | 0.761 | 0.730 | 0.759 | 0.763 | 0.745 | 0.752 |

**Table 2.** Segmentation results (DSC) of U-Net in 5-fold cross validation using the training image dataset.

|  | fold-1 | fold-2 | fold-3 | fold-4 | fold-5 | Average |
| --- | --- | --- | --- | --- | --- | --- |
| Patching | 0.731 | 0.712 | 0.752 | 0.704 | 0.723 | 0.725 |
| Resizing | 0.760 | 0.719 | 0.741 | 0.755 | 0.734 | 0.742 |

When applying the trained models on the 53 challenge testing cases, a bagging-type ensemble strategy was implemented to combine the outputs of these ten models to further improve the segmentation performance, achieving an average of DSC, precision and recall of 0.7318, 0.7851 and 0.7319 respectively, which ranked our method as the fourth place in the challenge.

### 5 Summary

In this work, we presented a fully automated segmentation model for head and neck tumor segmentation from PET and CT images. Our SA-Net replaces the long-range skip connections between the same scale in the vanilla U-Net with full-scale skip connections in order to make maximum use of feature maps in full scales for accurate segmentation. Attention mechanism is introduced to adaptively adjust the weights of each scale feature to emphasize the important scales while suppressing the less important ones. As compared to the vanilla U-Net structure with scale-wise skip connection and feature concatenation, the proposed scale attention block not only improved the segmentation performance by 2.25%, but also reduced the number of trainable parameters from 17.8M (U-Net) to 16.5M (SA-Net), which allowed it to achieve a top performance with limited GPU resource in this challenge. In addition, the proposed SA-Net can be easily extended to other segmentation tasks. Without bells and whistles, it has achieved the 3rd place in Brain Tumor Segmentation (BraTS) Challenge 2020^1^.

## Data Availability

The data used in this study is publicly available

https://www.aicrowd.com/challenges/hecktor

## Acknowledgment

This work is partially supported by a research grant from Varian Medical Systems (Palo Alto, CA, USA) and grant UL1TR001433 from the National Center for Advancing Translational Sciences, National Institutes of Health, USA.

## Footnotes

1 https://www.med.upenn.edu/cbica/brats2020/rankings.html

